# Prevalence and Risk Factors of Diabetes Mellitus in People over 40 Years Old in Rural Areas of Central Hunan Province

**DOI:** 10.1101/2020.05.23.20107995

**Authors:** Lan Li, Yuan Liao, Hao Xiao, Tiantian Wang, Jing Liu, Guijuan Zhou, Guanghua Sun, Kang Li, Fujin Huang, Weibin Feng, Jun Zhou

## Abstract

**Objective:** To explore the prevalence of diabetes mellitus in rural areas of central Hunan province, and to analyze the risk factors related to diabetes mellitus (DM).

**Methods:** A cross-sectional study was conducted by using questionnaires (gender, age, occupation, degree of education, exercise habits, smoke status, alcohol consumption, hypertension, cardiovascular disease, family history of diabetes), anthropometric measurements (height, weight, waist circumference, blood pressure) and biochemical indexes test (fasting blood-glucose, glycosylated hemoglobin, blood lipids). Villagers over 40 years old from a community in rural areas of central Hunan Province were investigated.

**Results:** A total of 410 clinical data were collected. The prevalence of diabetes mellitus in 410 (177 in male and 233 in female) villagers was 13.41%, including 13 males with a prevalence rate of 7.34% and 42 females with a prevalence rate of 15.16%. The prevalence of impaired fasting glucose (IFG) was 11.95%, 21 males (11.86% of males) and 28 females (10.11% of females). And the results of glycosylated hemoglobin test showed that 64% villagers with diabetes mellitus had hemoglobin A1c(HbA1c) above 6.5%. Univariate analysis suggested that gender, smoke status, alcohol consumption, family history of diabetes mellitus, hypercholesterolemia and hypertension were involved in diabetes mellitus (P<0.05). Multivariate logistic regression analysis showed that family history of diabetes (OR: 1.759; 95% CI: 1.010-3.065), hypercholesterolemia (OR: 3.819; 95% CI: 1.27-11.486) and hypertension (OR: 2.074; 95% CI: 1.130-3.809) were independent risk factors for diabetes mellitus, and the differences were statistically significant (P < 0.05).

**Conclusion:** The prevalence of diabetes mellitus in rural areas of central Hunan Province is higher. Family history of diabetes, hypercholesterolemia and hypertension are major risk factors for diabetes mellitus. The knowledge of diabetes should be strengthened. Related interventions should be given based on the diabetes epidemic status of local.

## Introduction

Diabetes mellitus, as well as hyperglycemia, is considered one of the most well-known disease affecting various body systems. The prevalence of Diabetes mellitus increases greatly with the aged tendency of population and change of life style, which severely threatens people’s health and affects the quality of life of adults even children^[1]^. Diabetes mellitus has been one of the biggest public health problems in the modern world, but people’s awareness of diabetes is not high. At present, domestic data on this aspect are mostly based on first-tier cities^[2-4]^, while there is rarely systematic and in-depth investigation on low-income areas such as rural areas in central Hunan province. The purposes of this study were to investigate the prevalence of diabetes and explore its risk factors among people over 40 years old in rural areas of central Hunan province, in order to enrich the prevalence of diabetes among residents in different regions in China and provide scientific evidence for the formulation of national health policies.

## Methods

### Study participants

In January 2019, a representative sample of the general participants aged ≥40 years was selected in order to investigate the prevalence and related risk factors in rural areas of central Hunan province. The study adopted the scheme of random sampling, and selected Jiao Shan Community in Hengyang city, Hunan province. All eligible permanent residents aged ≥40 years in each village were invited to participate in the study. The flow chart of this study was shown in **Fig. 1**. This study was subject to approval by the Ethics Committee of the First Affiliated Hospital of University of South China, Hengyang, Hunan. And clinical trial registration in Chinese Clinical Trial Registry Center, ChiCTR1800020006.All procedures were carried out in accordance with the ethical standards. Additionally, all participants, after being properly informed about the study procedures and potential risks, signed a free informed consent document.

**Fig. 1.**
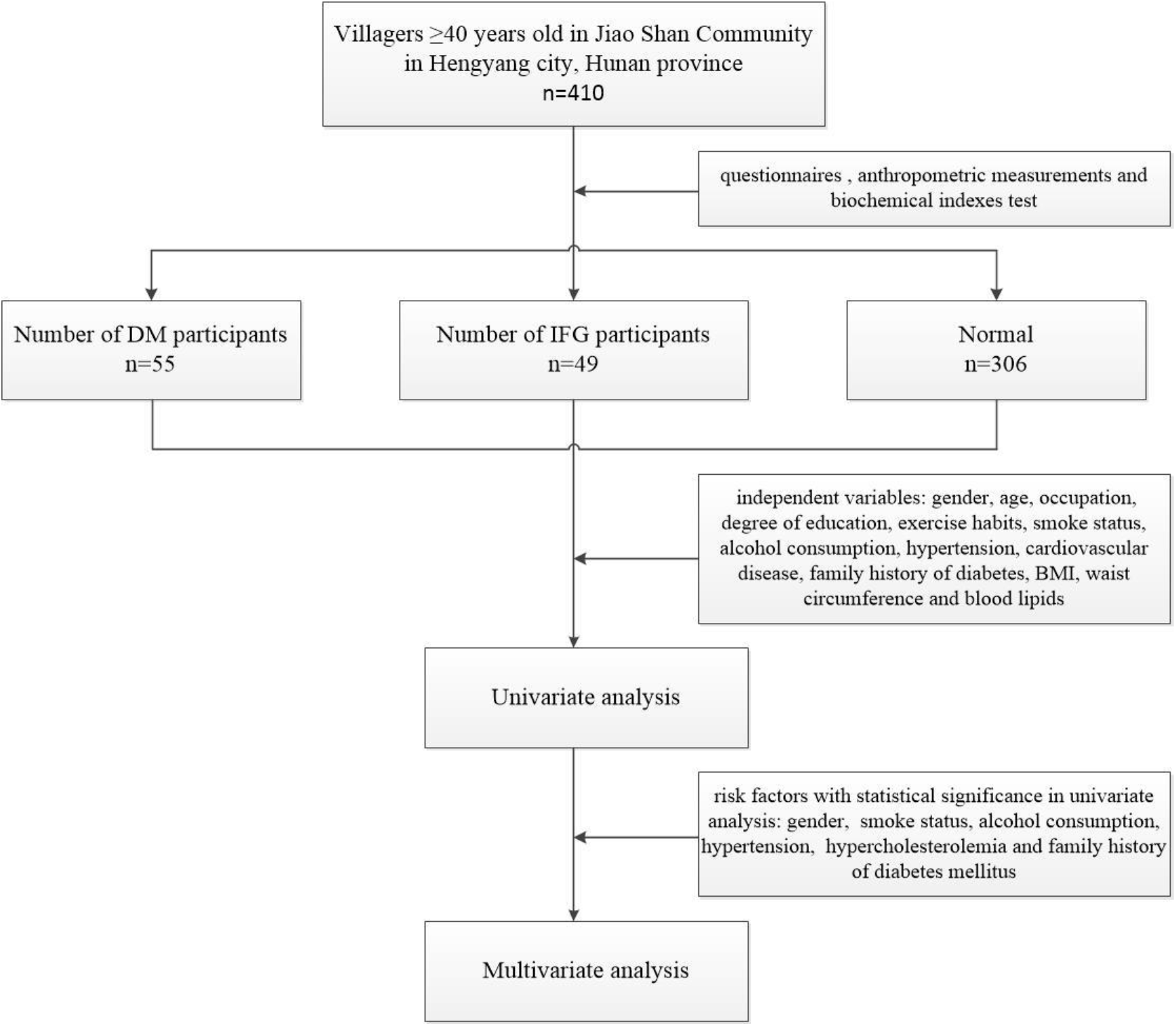
Process of the study

### Data collection and measurements

The methods of investigation include questionnaires (such as gender, age, occupation, degree of education, exercise habits, smoke status, alcohol consumption, hypertension, cardiovascular disease, family history of diabetes), anthropometric measurements (include height, weight, waist circumference, blood pressure) and biochemical indexes test (fasting blood-glucose, glycosylated hemoglobin, blood lipids and so on). The participants were instructed to take off their shoes and wear lightweight clothes When measuring height and weight. Height and waist circumference were accurate to 0.1cm and weight was accurate to 0.1kg. Waist circumference (WC) was measured at the umbilical level (to the nearest 0.1cm). The body mass index (BMI) was calculated as weight(kg)/height(m^2^).

Blood pressure was measured two times at one-minute intervals after at least five minutes of rest using a standardized electronic automatic sphygmomanometer (TSB-615B-T; Chaosi, Beijing, China). the average of the two blood pressure measurements was calculated at each examination.

Fasting venous blood was drawn and placed into a vacuum tube containing EDTA (Becton Dickinson, Franklin Lakes, NJ, USA) after fasting for at least 8 hours. Fasting plasma glucose (FPG) were measured using a blood glucose test paper (Sinocare Inc.).The correlation analyses of biochemical indices (total cholesterol, triglycerides, low-density lipoprotein cholesterol and high-density lipoprotein cholesterol) were identified by the Cadillac detector (SN:3222597,Indianapolis, USA).Centrifugation was done within 2 hours after collection and hemoglobin A1c was determined within 24 hours using a specific protein analyzer HP-AFS/3, Hebei, China).All laboratory equipment was calibrated and blinded duplicate samples were used.

### Definitions

According to the 2017 American Diabetes Association (ADA) guidelines diabetes diagnosis^[5]^ was that FPG≥7.0 mmol/L or using insulin or oral hypoglycaemic drugs or history of diabetes diagnosis. And IFG was defined FPG > 6.1mmol/L and < 7.0mmol/L. The diagnostic criteria for Dyslipidemia were based on the 2007 Chinese Guideline for Prevention and Treatment of Dyslipidemia in Adult Patients: total cholesterol (TC)≥6.2mmol/L; or triglycerides (TG)≥2.3mmol/L; or low-density lipoprotein cholesterol (LDL-C) ≥4.1mmol/L; or high-density lipoprotein cholesterol (HDL-C)< 1.0mmol/L was defined Dyslipidemia. We categorized participants into four groups according to their BMI: obese cases were BMI≥28.0kg/m^2^, overweight cases were BMI < 28.0Kg/m^2^ and BMI≥24.0Kg/m^2^; normal weight cases were BMI≤18.5Kg/m^2^ and BMI > 24.0Kg/m^2^ and underweight cases were BMI < 18.5Kg/m2^[6]^. Abdominal obesity^[7]^ was defined as a waist circumference in men≥ 90 cm, women≥ 80 cm. In this study, smoke status was defined as ever smokers (former or current smoking). Alcohol consumption (liquor)≥3 times per week and each ≥400g were defined heavy drinking; otherwise it’s a low drinking. Regular exercise was defined as exercising 3 to 5 times or more per week. Family history of diabetes was defined as having at least one parent or sibling with diabetes. What’s more, Hypertension was defined as systolic blood pressure ≥140 mmHg and/or diastolic blood pressure ≥90 mmHg, or taking antihypertensive drugs^[8]^. Coronary heart disease (CHD) was defined as previous history of coronary artery disease.

### Statistical analysis

All data were analyzed using the IBM SPSS Statistics for Windows version 22. A univariate analysis of risk factors was conducted with gender, age, occupation, degree of education, exercise habits, smoke status, alcohol consumption, hypertension, cardiovascular disease, family history of diabetes, BMI, waist circumference and blood lipid level as the independent variables. T-test was used for measurement data, chi-square(χ^2^) test was used for counting data, and Risk factors with statistical significance in univariate analysis were subjected to multivariate logistic regression analysis. P< 0.05 was considered statistically significant.

## Results

### Univariate analysis

A total of 410 participants [males:177(43.2%), females:233(56,8%)] were investigated in the study. The biochemical characteristics and risk factors of the studied group are shown in **Table 1**. The prevalence of diabetes mellitus in 410 villagers was 13.41%, including 13 males with a prevalence rate of 7.34% and 42 females with a prevalence rate of 15.16%. And the prevalence of IFG was 11.95%, 21 males (11.86% of males) and 28 females (10.11% of females) (**Table 2)**.

**Table 1.** The biochemical characteristics and risk factors of the population in rural areas of central Hunan Province

| Factor | Total | Men | Women | $\chi^2$ | P-value |
| --- | --- | --- | --- | --- | --- |
| **Age group(years)** | 410 |  |  | 6.69 | 0.154 |
| 40-49 | 33 | 14 | 19 |  |  |
| 50-59 | 134 | 54 | 80 |  |  |
| 60-69 | 185 | 83 | 102 |  |  |
| 70-79 | 42 | 19 | 23 |  |  |
| ≥80 | 16 | 7 | 9 |  |  |
| **BMI (kg/m<sup>2</sup>) \*** | 341 |  |  | 2.006 | 0.571 |
| ≥28 | 17 | 10 | 7 |  |  |
| 24-27.9 | 114 | 55 | 59 |  |  |
| 18.5-23.9 | 193 | 77 | 116 |  |  |
| <18.5 | 17 | 3 | 14 |  |  |
| **Education level \*** | 397 |  |  | 5.630 | 0.131 |
| pre-school or Primary school | 269 | 95 | 174 |  |  |
| Secondary school | 107 | 60 | 47 |  |  |
| Tertiary education | 19 | 14 | 5 |  |  |
| **Occupation\*** | 399 |  |  | 0.207 | 0.649 |
| Crop and Pasture Production | 330 | 131 | 199 |  |  |
| other | 69 | 43 | 26 |  |  |
| **Smoke status \*** | 408 |  |  | 6.958 | 0.008 |
| yes | 121 | 116 |  |  |  |
| no | 287 | 59 |  |  |  |
| **Alcohol consumption \*** | 407 |  |  | 7.615 | 0.022 |
| no | 283 | 87 | 196 |  |  |
| light | 91 | 56 | 35 |  |  |
| heavy | 33 | 31 | 2 |  |  |
| **Exercise \*** | *403* |  |  | 0.045 | 0.832 |
| often | 305 | 133 | 172 |  |  |
| rarely | 98 | 41 | 57 |  |  |
| **Waist circumference (cm) \*** | *400* |  |  | 0.246 | 0.620 |
| ≥90 in men, ≥80 in women | 129 | 38 | 91 |  |  |
| <90 in men, <80 in women | 271 | 134 | 137 |  |  |
| **Hypertriglyceridemia\*** | *395* |  |  | 1.603 | 0.206 |
| yes | 99 | 46 | 53 |  |  |
| no | 296 | 124 | 172 |  |  |
| **Hypercholesterolemia\*** | *395* |  |  | 8.235 | 0.004 |
| yes | 16 | 4 | 12 |  |  |
| no | 379 | 166 | 213 |  |  |
| **High LDL cholesterolemia\*** | *365* |  |  | 1.003 | 0.317 |
| yes | 13 | 7 | 6 |  |  |
| no | 352 | 149 | 203 |  |  |
| **Low HDL cholesterolemia\*** | *395* |  |  | 0.170 | 0.680 |
| yes | 83 | 47 | 36 |  |  |
| no | 312 | 123 | 189 |  |  |
| **Hypertension** | *410* |  |  | 6.431 | 0.11 |
| yes | 160 | 72 | 88 |  |  |
| no | 250 | 105 | 145 |  |  |
| **Coronary heart disease \*** | *348* |  |  | 0.073 | 0.787 |
| yes | 32 | 14 | 18 |  |  |
| no | 316 | 136 | 180 |  |  |
| **Family history of diabetes** | *410* |  |  | 16.5 | 0.000 |
| yes | 33 | 11 | 22 |  |  |
| no | 363 | 160 | 203 |  |  |

\* missing values

**Table 2.** The prevalence of diabetes mellitus and IFG and their HbA1c level among people over 40 years old in rural areas of central Hunan province

| Variable | DM |  | IFG |  |
| --- | --- | --- | --- | --- |
|  | Number of DM participants | Prevalence (%) | Number of IFG participants | Prevalence (%) |
| Men(n=177) | 13 | 7.34 | 21 | 11.86 |
| Women(n=233) | 42 | 15.16 | 28 | 10.11 |
| Total(n=410) | 55 | 13.41 | 49 | 11.95 |
| HbA1c>6.5% | 35 | 64 | 3 | 6.1 |

It’s known that hemoglobin A1c above 6.5 indicates poor glycemic control in the last 2-3 months ^[9]^. In our study, the results of glycosylated hemoglobin test showed that 35 patients with diabetes mellitus had no blood glucose control in the past 2-3 months, and the rate of non-compliance reached 64% (**Table 2**).

Hypercholesterolemia in dyslipidemia is major risk factors for diabetes mellitus in both men and women **(Table 1)**. Furthermore, univariate analysis suggested that gender, smoke status, alcohol consumption, family history of diabetes mellitus, hypercholesterolemia and hypertension were involved in diabetes mellitus (P<0.05) **(Table 1)**. The prevalence of diabetes generally increased with age, which was more noticeable in females than in males (**Table 2 and Fig. 2**). The prevalence of diabetes in 40-49 years old was less than that of IFG, while the prevalence of IFG was lower than that of diabetes in over 50 years old (**Fig. 2**).

**Fig. 2.**
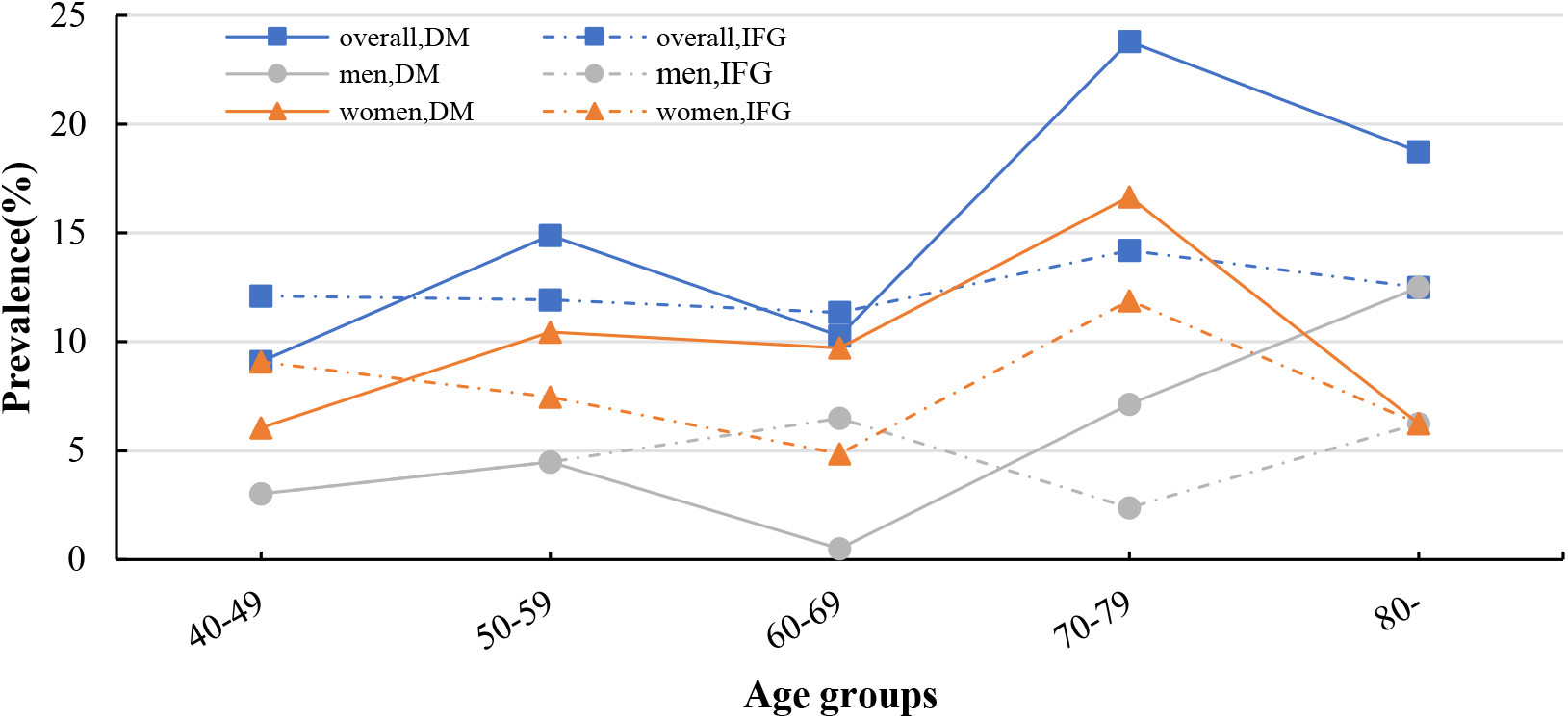
The prevalence of diabetes mellitus and IFG among villagers of different ages

### Multivariate analysis

Subsequently, that gender, smoke status, alcohol consumption, family history of diabetes mellitus, hypercholesterolemia and hypertension had statistical significance in univariate analysis was incorporated into the multivariate analysis. The result of multivariate logistic regression analysis is showed that family history of diabetes (OR: 1. 759; 95% CI: 1.010-3.065), hypercholesterolemia (OR: 3.819; 95% CI: 1.270-11.486) and hypertension (OR: 2.074; 95% CI: 1.130-3.809) were independent risk factors for diabetes mellitus, and the differences were statistically significant (P < 0.05) **(Table 3)**.

**Table 3.** Multivariate analysis for diabetes mellitus risk factors

| Variable | Odd ratio | 95%CI | P-value |
| --- | --- | --- | --- |
| Gender | 0.639 | 0.261-1.566 | 0.328 |
| Smoke status | 0.458 | 0.147-1.424 | 0.177 |
| Alcohol consumption | 0.646 | 0.333-1.256 | 0.198 |
| Hypercholesterolemia | 3.819 | 1.27-11.486 | 0.017 |
| Hypertension | 2.074 | 1.130-3.809 | 0.019 |
| Family history of diabetes | 1.759 | 1.010-3.056 | 0.046 |

## Discussion

Diabetes mellitus poses serious public health problems, closely related to a variety of diseases, such as cerebral infarction and coronary heart disease, affecting individuals and society in terms of life quality and economic burden^[10-12]^. It is reported that women may exercise less than men, and a series of physiological changes associated with the relatively lower level of sex hormones after menopause in women affect blood glucose levels in the body^[13]^.

In this study, the prevalence of diabetes generally increased with age, which was more noticeable in females than in males. This study was a community-based, cross-sectional survey carried out in rural areas of central Hunan Province, China. And the prevalence of the areas is 13.41%, which is higher than Liaoning and Sichuan province^[4, 14, 15]^. Recently, more and more people began to occupied in sedentary lifestyle instead of manual labor, and the risk of diabetes increases with lack of physical activity^[16]^. Adequate physical activity can increase insulin sensitivity, and is important to preventing the development of many chronic diseases such as diabetes, hypertension and obesity^[17, 18]^. The presented study shows that their occupation is not a risk factor for diabetes, which is related to the fact that most villagers have been engaged in crop and pasture production (moderate to severe physical labor) for a long time, indicating that physical labor can reduce the occurrence of diabetes in rural China.

It is known that IFG is pre-diabetes^[19]^, and the prevalence of IFG in this study was 11.95%, which is slightly lower than diabetes mellitus. If they are not taken seriously, it is easy for them to further develop into diabetes, which in turn increases the prevalence of diabetes. Actually, most villagers have few symptoms that may not be recognized, are not aware of their pre-diabetes, and do not control diet and strengthen exercise, which may be one of the reasons that the prevalence of diabetes in this region is higher than in other regions. In addition, diabetes control in these diabetic villagers is not adequate as showed by high rates of poor control of HbA1c, because of the low education level of the villagers, the weak awareness of diabetes prevention and control, and the poor treatment compliance.

Some, but not all, studies^[20, 21]^ have shown that smoking is associated with an increase in systemic insulin resistance. Maddatu et al.^[22]^ also showed that compared with non-smokers, long-term smokers can inhibit insulin signaling pathways, strengthen fat infiltration in muscles, reduce adiponectin levels, increase retinol binding protein 4 and increase insulin resistance in skeletal muscles. In this research, alcohol consumption increase the risk of diabetes mellitus, which is different from one study that reported that light alcohol consumption can reduces the risk of diabetes^[23]^. Multicenter studies with large sample size are needed for further analysis and verification.

Both univariate and multivariate analyses suggested that hypercholesterolemia increased the risk of diabetes. Sabag et al.^[24]^ reported that exercise can effectively improve obesity and abnormal lipid metabolism in Type 2 diabetes mellitus(T2DM) patients. Moreover, residents with positive family history of diabetes had a higher prevalence than those without family history of diabetes, which is related to family dietary habits, lifestyle similarity and genetic factors.

Increased adipokine secretion, lipid deposition, inflammatory response, oxidative stress, and metabolic abnormalities associated with insulin resistance are the common pathophysiological basis of obesity, diabetes, and coronary heart disease^[25]^. Increased adipose tissue accelerates oxidative stress response in obese patients, and the accumulation of intracellular reactive oxygen leads to decreased insulin sensitivity through protein kinase C, mitokinase-activated protein kinase p38 and c-jun amino-terminal kinase^[26, 27]^. A series of metabolic disorders such as hyperglycemia and increased free fatty acids which caused by decreased insulin sensitivity has a direct toxic effect on cells^[28]^, promoting the occurrence of diabetes.

There are common risk factors for diabetes and hypertension, which may act on both diseases^[29]^. Diabetes is a risk factor for hypertension, and it highlighted separately in guidelines on hypertension in China. As a risk factor for diabetes, hypertension has a great influence on the occurrence and development of diabetes, closely to abnormal blood pressure regulation, endothelial dysfunction and lipid metabolism disorder^[30]^.

The study has great significance to the formation of diabetes prevention and control strategies in central Hunan province and even the country, but there are also some deficiencies: First, some villagers over 40 years old are not included in the survey because they are out of town or far away from the survey site, and the study subjects may have gender deviation. Second, although type 2 diabetes is the dominant type in adults over the age of 40, but type 1 and type 2 diabetes were not classified in this study. Third, the risk factors in this study did not include recognized risk factors such as obesity and exercise, which may be related to insufficient sample and the erroneous definition of exercise. Additionally, dietary habits were not included in this study because of the unreasonable questionnaire design. In the future, more large-sample size is needed to obtain more scientific evidences.

## Conclusion

The prevalence of diabetes mellitus in rural areas of central Hunan Province is higher, and most diabetic villagers have failed to achieve glycemic control in the past 2-3 months. Family history of diabetes, hypercholesterolemia and hypertension are major risk factors for diabetes mellitus. The knowledge of diabetes should be strengthened. Related interventions should be given based on the diabetes epidemic status of local.

## Data Availability

The data used to support the findings of this study are included within the article

## Acknowledgements

We would like to thank Shiju Chen MD and Chengxiao Fu MD for their assistance with language editing of this manuscript. And this study was supported by the Natural Science Foundation of Hunan Province (2018JJ2358).

## Notes

### Competing Interest Statement

The authors have declared no competing interest.

### Author Declarations

This study was subject to approval by the Ethics Committee of the First Affiliated Hospital of University of South China, Hengyang, Hunan.

